# A mixed-methods feasibility study of an educational intervention to operationalize recommendations for community-academic genomics research partnerships

**DOI:** 10.64898/2026.08.14.26360484

**Authors:** Letonia Copeland-Hardin, Katalina Salas, Michael V. Rodriguez, Kelsie Huff, Brandi M. White, Marcia Tan

## Abstract

**Background:** Scientific mistrust contributes to lower participation and underrepresentation of Black Americans in genomics studies limiting understanding of how genomic variation and environmental exposures influence health disparities. Community-engaged research requires rebuilding scientific trust; however, there is a need for practical models that operationalize guidance for researchers without community-engaged research training. Therefore, we developed an educational intervention for genomics researchers initiating partnerships with Black American communities. Our intervention creates a bidirectional teaching environment that allows potential community and academic partners to discuss each stakeholder’s area of expertise, partnership perspectives and needs, while exploring modules related to genomics research and community-academic partnership. We report a novel and structured approach for prospective academic and community partners to mutually orient one another and assess partnership practicability.

**Methods:** Ten participants, recruited through our community partner’s established channels, attended a four-hour workshop containing nine interactive modules about scientific mistrust, research safeguards, community-engaged research, genomics, and research for community-defined goals. We also experimented with humor to enhance engagement and trust. We used mixed-methods, single-arm research design and assessed feasibility through recruitment success and retention. Using inductive rapid thematic analyses, we assessed participant responses to eight workshop prompts. Preliminary quantitative data, used for descriptive purposes due to low sample size, were analyzed from pre- and post-Likert scale surveys assessing associations between the intervention and four domains, including scientific trust.

**Results:** All 10 enrolled participants completed the study, meeting a priori criteria of 100% for recruitment and show rate. Engagement was strongest during early workshop modules and declined in later modules. Survey data completeness was limited by missing responses. Survey instrument design limitations were identified for modification in future studies. Participants articulated expectations for partnership that aligned with community-based participatory research principles.

**Conclusions:** The intervention is feasible to deliver as a bidirectional educational experience in partnership with a community organization. The strongest implementation refinements needed were workshop duration, module prioritization, and survey instrument design, particularly the trust domain. A community advisory board is co-developing modules and refining the intervention to evaluate in a pilot version of the study with a larger sample.

## BACKGROUND

Genomic research has historically reflected and perpetuated inequities in participation and benefit, with individuals of European ancestry disproportionately represented in studies, while racially minoritized populations, including Black Americans, remain significantly underrepresented [1–5]. Underrepresentation limits the scientific community’s understanding of how genomic variation impacts adverse health outcomes within and across diverse populations [5–8]. Population-level studies identifying genomic features and environmental factors driving health disparities that negatively and disproportionately affect Black Americas require biospecimens from Black Americans to support the development of pharmacological and environmental-related interventions that improve health outcomes. Therefore, eliminating health disparities requires greater inclusion of underrepresented groups to fully characterize the impact of the spectrum of human genetic diversity on health and disease outcomes [5, 9].

A complex combination of historical, cultural, and institutional factors contributes to the European bias in genomic and environmental omics studies, including lower participation of Black Americans [10, 11]. Although there have been attempts to improve diversity in biomedical research, underrepresentation persists partially because of scientific and medical mistrust [12]. Mistrust of research institutions stems from historical transgressions, ongoing discrimination in healthcare, lack of transparency, and insufficient public engagement by scientific researchers [12–15]. In addition, behaviors considered extractive, such as interacting with communities solely to collect biospecimens and information without considering mutual benefit or respecting community-defined priorities, may reduce the trustworthiness of scientists [16].

Community-based participatory research (CBPR), which positions the community partner as an equitable stakeholder at each stage of the research process, involves building trust between communities and scientists [17–19]. CBPR promotes collaborative engagement from the ideation of research questions through the dissemination of results, while encouraging bidirectional learning and valuing the expertise of both community and academic partners. While the implementation of CBPR varies according to stakeholder preferences, research scope, and funding requirements, progress depends fundamentally on building and maintaining trust within community-academic partnerships [20].

The American Society of Human Genetics (ASHG) published guidance indicating the importance of community-engaged approaches in genomics research [21]. Models, such as the Community Engagement Studio (CE Studio) exist for obtaining input from community stakeholders on existing research projects [22]. Yet, there remains a need for practical models for genomics researchers, addressing the earlier phase of initiating a community-academic partnership, particularly ones that build researchers’ capacity for direct community engagement. Despite the reported benefits of CBPR in improving the diversity and translational potential of genomics research, community-engaged approaches are not traditionally incorporated into graduate-level training for genomics researchers [23]. Additionally, many researchers are trained in investigator-led methods that position them as leaders instead of collaborators with community stakeholders. As a result, opportunities for mutual benefit are overlooked and community perspectives may not be part of the research design. Moreover, some researchers with community-engaged research training, may have relied on university administrators and staff to establish community partnerships, which can prevent trainees from developing the skills needed to independently build new community-academic partnerships. Researchers without prior community-engaged research training may benefit from practical tools focused on trust-building, co-learning, and reducing information asymmetry that are useful to researchers and communities entering partnerships.

Our intervention offers genomics researchers a structured framework to learn community-defined priorities and present their research to prospective partners, while giving community partners access to information on participant safeguards, a framework for conveying their terms of engagement, and a means of evaluating a prospective academic partner’s research focus. The primary objective of the reported study is to assess the feasibility of this intervention through recruitment success, and retention. Level of engagement was also examined. For descriptive purposes, we generated preliminary data examining the intervention’s impact on trust in scientists, willingness to engage with researchers, viewing research as useful for supporting community goals, and comfort with humor throughout the intervention. This feasibility study evaluated whether the workshop could be executed as planned and identified optimizations necessary for our planned pilot study with a larger sample of participants.

## METHODS

### Study Design and Community Partner

We conducted an uncontrolled, single-arm, mixed-methods feasibility study. Our community partner is People for Community Recovery (PCR), a registered 501(c)(3) nonprofit grassroots organization based in Chicago, IL that advocates for the environmental justice and health needs of residents of Altgeld Gardens and Phillip Murray Homes, a public housing community. For decades, PCR has organized community-based health initiatives and events and worked with elected officials, and has established its members as recognized leaders within the Altgeld Gardens community. Altgeld Gardens is situated in Riverdale, one of Chicago’s 77 official community areas, and is part of “the toxic doughnut” due to being surrounded by numerous polluting entities [24]. Riverdale, and therefore Altgeld Gardens, exhibits significantly higher diagnosis rates of certain cancer types relative to the broader Chicago population, and the community is exposed to the second-highest levels of ambient air pollution among Chicago neighborhoods [25–27]. PCR has long been interested in characterizing relationships between pollution and cancer health outcomes in this community; however, establishing community-academic partnerships and securing the support and engagement of other Altgeld Gardens residents is essential for conducting population-level studies, including genomics studies requiring biospecimens.

### Participants

An email and flyer were shared with a PCR representative to recruit participants. Participants (N=10) actively participated in a single, four-hour workshop. To maximize obtaining the most meaningful insights within a small sample size, and increase the likelihood of community perspectives informing the future larger study, participants were required to have lived experience in the Altgeld Gardens community. Previous residents were required to hold leadership positions within or be members of PCR. Half of the participants were PCR members familiar with the content shared in the workshop. Their inclusion was strategic to support interactive discussion during workshop prompts; PCR members have community knowledge to answer prompts and model engagement for less-experienced participants. In addition, we believed that their presence would ensure that community perspectives were prioritized during prompt discussions and provided mechanisms for conveying their perspectives and experiences to the researcher leading the study, which placed the researcher in a learner role. Additional eligibility requirements included being at least 18 years of age, and identifying as Black American.

### Intervention Development

Between January 2025 and June 2025, the curriculum was developed by “The Neighborhood Scientists,” a group of postdoctoral scholars and stand-up comics interested in civic science and community-engaged research. We used Kern’s Six Step Approach to Curriculum Development, a systematic framework for designing medical education programs while creating and organizing the modules [28]. Our goals included enhancing trust in science, improving willingness to engage with scientists, and viewing research as a tool for achieving community-level environmental justice and health goals. Each goal mapped to three of our four survey domains used to assess the intervention (see Survey Instrument). To accomplish these goals we developed a workshop as an education intervention aimed to 1) improve transparency about research processes, participant protections, and the expectations and responsibilities in equitable academic-community partnerships (Table 1, modules 1-5) 2) improve foundational scientific knowledge about genetics and pollutant-driven mechanisms related to cancer (Table 1, modules 7 and 9) 3) facilitate bidirectional learning in a co-teaching environment that integrates community perspectives and scientific knowledge and 4) provide practical information that community stakeholders can use strategically to leverage research to advance community priorities (Table 1, modules 6, 8, and 9). Finally, the last module of the intervention involved reviewing a peer-reviewed study that bridged the community’s environmental health and cancer interests with the researchers’ expertise, as an example of what a potential future study could entail. [29].

**Table 1:** Curriculum Structure by Module.

| Module | Title | Description |
| --- | --- | --- |
| 1 | Roots of Medical and Scientific Mistrust | Introduces ethical and legal requirements, guidelines, and policies in research aimed at protecting research participants; include the Nuremberg Code, Declaration of Helsinki, National Research Act, Belmont Report (informed consent), the Common Rule, and continuous ethics training for researchers |
| 2 | Safeguards to Protect Research Participants | Summarizes key rights for research participants including being informed, freely choosing participation, privacy, safety, asking questions, refusing or withdrawing participation, receiving a copy of the consent form, and information on reporting violations to Institutional Review Boards |
| 3 | Research Participant Rights | Involves comparing investigator-led ("helicopter") and community-engaged research approaches; involves discussion of the continuum of community engagement in research |
| 4 | "Helicopter Research" vs Community-Engaged Research | Covers fundamental characteristics of a trustworthy academic partner, including 1) openness to sharing research goals, methods, and risks (transparency) 2) valuing input, feedback, and expertise from community partners and practicing cultural humility (respect) 3) frequent and honest discussion about findings and progress (clear and open communication) and 4) commitment to ensuring research is conducted ethically (ethical standards) |
| 5 | Selecting Academic Partners | Provides an example of epidemiological research supporting policy and law changes; uses cigarette smoking as a case study |
| 6<sup>1</sup> | Scientific Research Impacting Public Policy and Law | Introduces concepts related to the genome, epigenome, transcriptome, and proteome |
| 7<sup>1</sup> | Genetics 101 | Overview of the peer-review process, and reasons publishing in peer-reviewed journals is important for establishing credibility to research intended to support community-prioritized policy goals |
| 8<sup>1</sup> | The Peer-Review Process | Review of a peer-reviewed research study that provides an example of a research question that may align with community research goals; the study investigates how pollution impacts gene expression and cancer. |
| 9 | Review of Published Genetic and Environmental Research Study | Short-term transcriptome and microRNAs responses to exposure to different air pollutants in two population studies |

<sup>1</sup> Module included the deliberate use of humor.

### Integration of Humor

Our broader trust-building, and engagement strategy involved the incorporation of humor as a pedagogical tool to make scientific concepts more accessible to a lay audience. Humor was strategically avoided during more serious and sensitive modules concerning scientific and medical mistrust, ethics, and forming academic-community partnerships to minimize the risk of further reducing mistrust of scientists and undermining the integrity of the content. Humor has been used as a pedagogical tool, can help learners retain information taught in lectures, and improves engagement in educational settings [30–37]. In addition, humor was grounded in published research suggesting that merging anthropomorphism satire can increase trustworthiness and likability of scientists [32].

### Study Procedure

The workshop was structured into seven distinct parts over a 4-hour period. The session began with a group informed consent process, during which the facilitator reviewed the consent form collectively and participants completed their forms privately. Following a brief break to ensure all were comfortable proceeding, the workshop continued with an introduction to the study and an icebreaker activity where participants created humorous mad scientist names and shared what they hoped to learn. Participants were then given an overview of the workshop agenda and completed pre-intervention surveys before moving into the main curriculum modules and workshop prompts. The intervention concluded with the completion of post-intervention surveys.

### Data Collection

#### Survey instrument

Participants completed paper-based pre- and post-intervention surveys in person immediately before and after the workshop. The two surveys were identical in content, except demographic information was collected at the end of the post-intervention survey. The instrument comprised 21 Likert-scale items grouped into four domains including comfort with humor in scientific communication (Humor), willingness to engage with scientists (Engagement), perception of research as a tool for environmental justice (Impact), and trust in scientists (Trust). Items 11–21 were adapted from the Medical Mistrust Index (MMI) [38], revised to focus on scientists rather than medical professionals. The adapted items were not psychometrically validated in this population. An open-ended text box was included on both surveys to allow participants to share unprompted feedback.

#### Workshop prompts

Ten brief, open-ended prompts were developed by the research team to encourage reflection and dialogue. Each prompt was displayed on a slide, with one to two prompts used to initiate individual workshop modules. Participants worked in pairs to record responses on Post-it notes, which were displayed collectively on a large board before being shared with the full group. Due to time constraints, two prompts were not delivered. Therefore, eight prompts were delivered and associated responses were collected at the close of each activity and transcribed verbatim for analysis.

#### Feasibility

The objective of this feasibility study was to evaluate whether the workshop could be delivered as designed and to identify modifications required prior to a subsequent pilot study. Feasibility data were collected to assess recruitment, show rate, data completeness, and protocol adherence. Our community partner identified and enrolled participants to a prespecified target sample size of 10. Attendance was recorded at the beginning of the workshop and used to calculate the show rate. Survey data completeness was assessed by examining pre- and post-intervention survey instruments for missing responses within each domain. Protocol adherence was defined as delivery of all workshop modules and prompts as specified in the protocol and deviations were documented by the facilitator following the session.

### Data Analysis

RStudio was used to analyze quantitative data, and create tables and figure 1.

**Figure 1.**
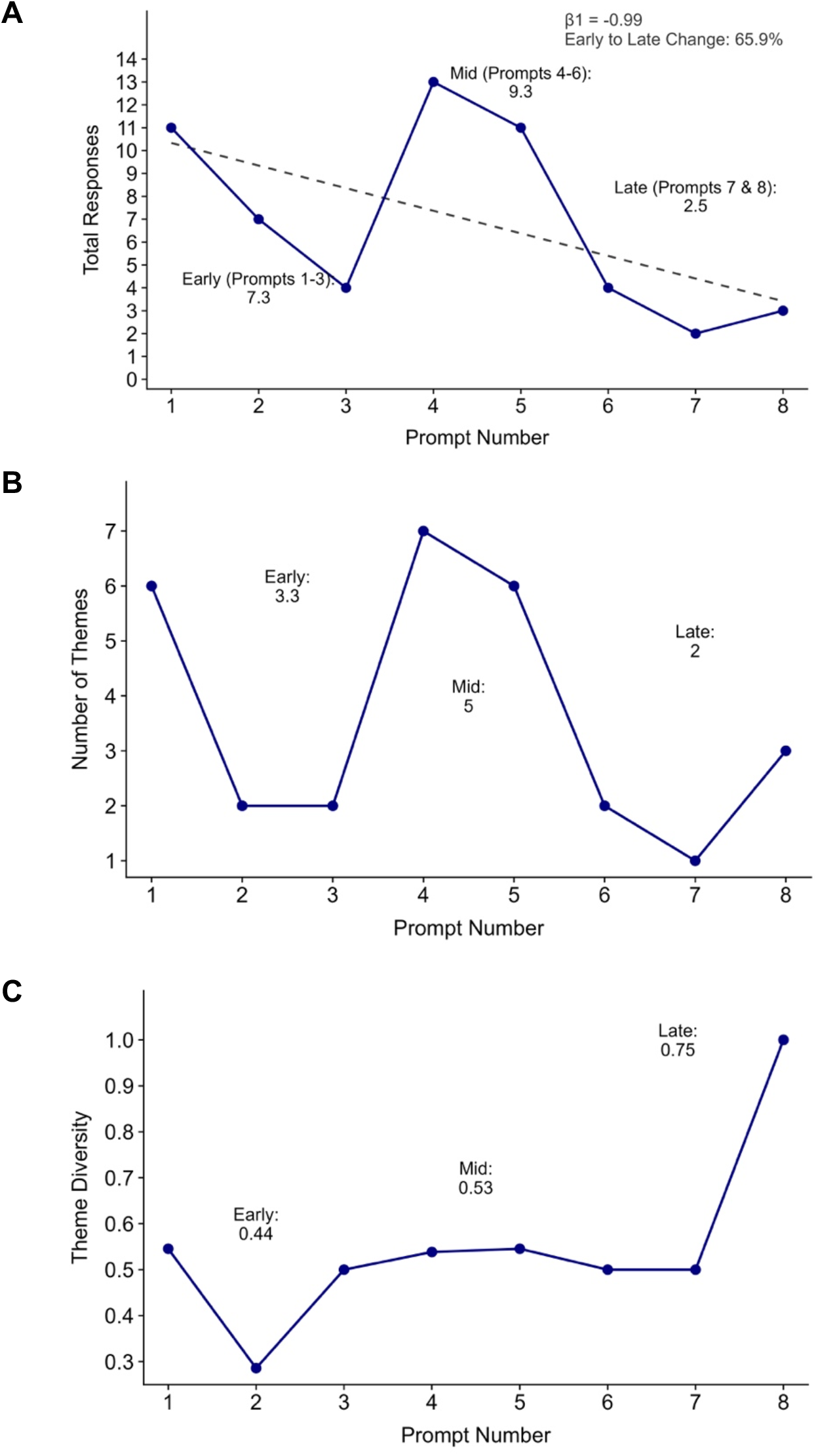
Engagement, unique themes, and theme diversity across prompts. (A) Engagement over time. Total responses submitted per prompt, plotted against prompt number with a fitted linear regression line. The slope (β_1_) is annotated on the panel. Relative prompt position serves as a proxy for time. The percentage change in responses from early to late is displayed. (B) Unique themes over time. Number of unique themes identified per prompt, plotted against prompt number. (C) Theme diversity over time. Ratio of total unique themes to total responses for each prompt, representing the average number of distinct themes contributed per response, plotted against prompt number. Mean response counts, mean theme counts, and mean theme diversity are shown for the early (prompts 1–3), mid (prompts 4–6), and late (prompts 7–8) prompt periods. The response and theme count for each prompt is reported in Table 2.

**Table 2:**
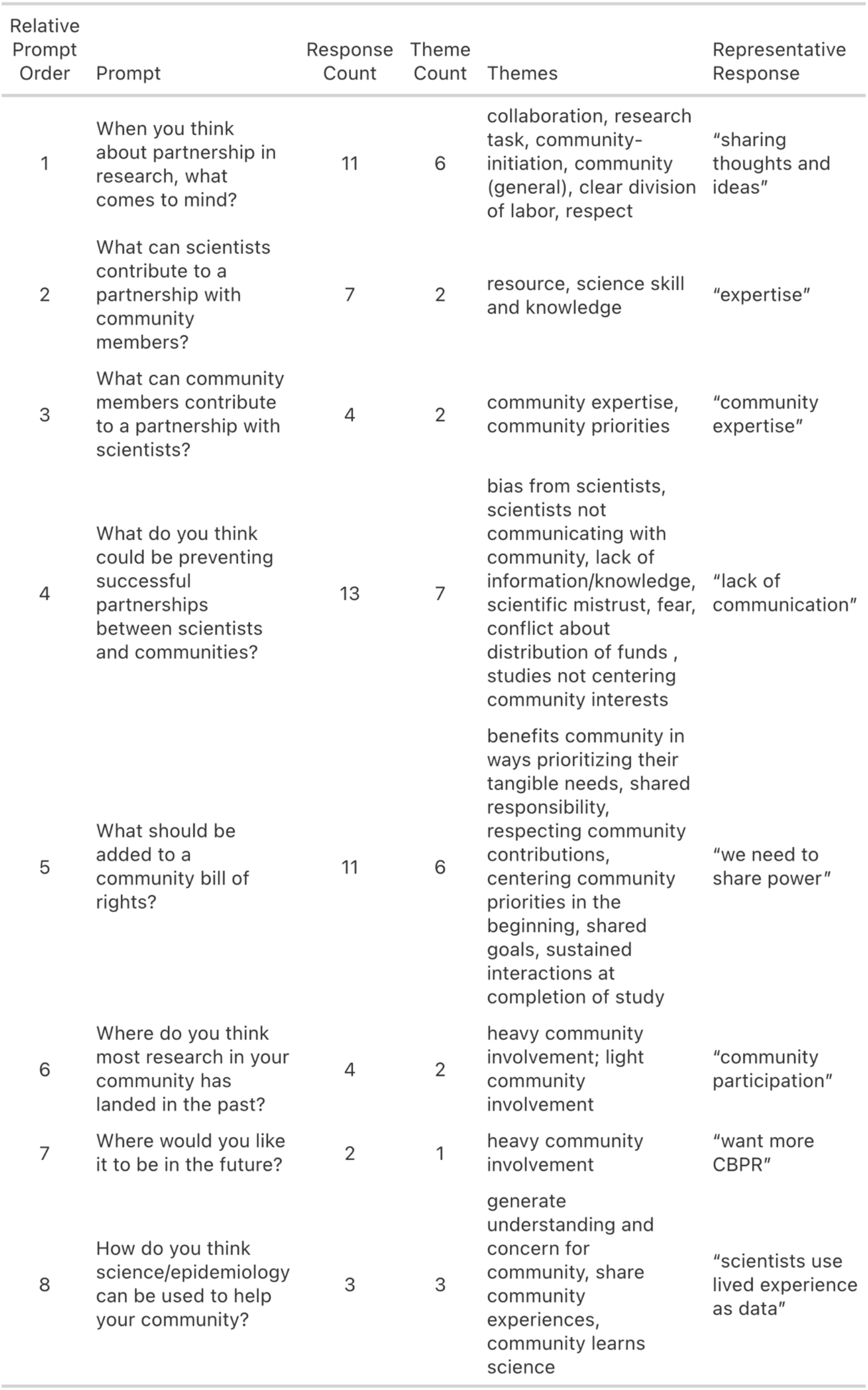
Summary of Result from Separate, Rapid Inductive Thematic Analyses.

| Relative Prompt Order | Prompt | Response Count | Theme Count | Themes | Representative Response |
| --- | --- | --- | --- | --- | --- |
| 1 | When you think about partnership in research, what comes to mind? | 11 | 6 | collaboration, research task, community-initiation, community (general), clear division of labor, respect | "sharing thoughts and ideas" |
| 2 | What can scientists contribute to a partnership with community members? | 7 | 2 | resource, science skill and knowledge | "expertise" |
| 3 | What can community members contribute to a partnership with scientists? | 4 | 2 | community expertise, community priorities | "community expertise" |
| 4 | What do you think could be preventing successful partnerships between scientists and communities? | 13 | 7 | bias from scientists, scientists not communicating with community, lack of information/knowledge, scientific mistrust, fear, conflict about distribution of funds, studies not centering community interests | "lack of communication" |
| 5 | What should be added to a community bill of rights? | 11 | 6 | benefits community in ways prioritizing their tangible needs, shared responsibility, respecting community contributions, centering community priorities in the beginning, shared goals, sustained interactions at completion of study | "we need to share power" |
| 6 | Where do you think most research in your community has landed in the past? | 4 | 2 | heavy community involvement; light community involvement | "community participation" |
| 7 | Where would you like it to be in the future? | 2 | 1 | heavy community involvement | "want more CBPR" |
| 8 | How do you think science/epidemiology can be used to help your community? | 3 | 3 | generate understanding and concern for community, share community experiences, community learns science | "scientists use lived experience as data" |

#### Feasibility assessment

A combination of quantitative and qualitative analyses was completed to assess feasibility. A priori feasibility benchmarks were established at 100% for quantitative metrics such as recruitment (N=10 participants), show rate, and data completeness. We had a goal of 0% attrition. Recruitment was calculated as the percentage of the target sample enrolled, while attendance was used to calculate show rate as the number of participants in attendance divided by the number of participants enrolled. Survey data completeness was calculated as the number of participants with complete responses to all items within each domain for both pre- and post-survey instruments. Protocol adherence, or implementation of workshop prompts, was qualitatively assessed by comparing workshop delivery against the protocol. Deviations were noted, including the number of prompts not delivered and the rationale.

#### Survey domain-level changes

To understand the preliminary impact of the intervention on the four survey domains, domain-level differences were compared using Likert-scale responses between the pre- and post-survey instruments. Survey data were analyzed quantitatively. Given the small sample size and incomplete responses on some items, survey results are reported descriptively to summarize observed changes following the intervention. Inferential statistical tests were not performed, and the study was not powered for hypothesis testing. Likert-scale responses for the 21 survey items were coded on a five-point scale, with reverse scoring applied to negatively worded items for consistent directional interpretation across items. Although individual Likert items are ordinal, the responses were treated as continuous because of the five-point scale structure and the use of domain mean scores. For each participant, domain mean scores were calculated as the mean of items within each domain. A complete case approach was used at the domain level. Therefore, participants missing a response to any item within a given domain at either the pre- or post-intervention assessment were excluded from analyses for that specific domain. Domain-level change was evaluated by comparing domain mean scores and standard deviations before and after the intervention.

#### Inductive thematic analysis

Participant responses to each of the workshop prompts were analyzed using inductive rapid thematic analysis. Responses for each prompt were analyzed independently because prompts addressed distinct topics. For each prompt, each response was initially coded, then related codes were grouped into emerging themes based on patterns. These themes represent the collective perspectives, experiences, and needs of participants related to research and academic-community partnerships. Coding was conducted by a single researcher and inter-coder reliability was not evaluated.

#### Participant engagement

Mixed methods were utilized to examine participant engagement throughout the intervention. Measures derived from the rapid, inductive thematic analysis were used as indicators of engagement including the total number of responses per prompt, and number of emerging themes per prompt. Higher response and theme counts were interpreted as indicating stronger engagement and a wider range of ideas. To examine change in engagement over the course of the workshop, the eight delivered prompts were divided into early, middle, and late periods comprising prompts one through three, four through six, and seven and eight, respectively. For each period, the mean number of responses and mean number of distinct thematic categories were calculated across the prompts in that period. In addition, a simple linear regression of response count on prompt order was fit, with prompt order serving as a proxy for time elapsed during the workshop. Given the small number of prompts (n = 8 prompts), regression results are interpreted descriptively as exploratory. The associated p-value is reported for transparency and is not interpreted as a test of statistical significance.

#### Theme diversity

The theme diversity ratio was created to examine the variety of responses reported by participants as a group to each prompt. It is calculated as the number of unique themes divided by the total number of responses per prompt. The ratio is dependent on response count, making higher values a potential reflection of fewer total responses rather than greater variety of ideas. Therefore, lower theme diversity may indicate lower engagement.

#### Participant interest

Free-text responses from the pre- and post-intervention surveys were reviewed by the research team for comments relating to interest in specific topics, modules, and the workshop overall. No formal analysis was applied.

### Ethics Approval and Consent to Participate

The University of Chicago Institutional Review Board approved of our study protocol (IRB25-0476). Informed consent was obtained from each participant.

## RESULTS

All participants self-identified as Black or African American, and the most common age group was 25-34 (40%). Educational levels varied, with most having some college (50%) or a high school diploma/GED (30%). The majority were current residents of Altgeld Gardens (70%), and the duration of residency for current and former residents was relatively evenly distributed with 30% having lived there for 1–5 years, 6–10 years, or at least 20 years. Half of the participants currently work or volunteer for PCR. Additionally, 50% had no prior experience attending workshops or educational programs about scientific research, while half of the participants had never interacted with a scientist. However, 50% had previously volunteered as research participants in other studies.

### Feasibility Outcomes

Feasibility measures were highest for recruitment, show rate, attrition and resource availability, with limitations related to implementation of workshop prompts, and data completeness. PCR successfully recruited 10 participants, resulting in a 100% show rate, with zero attrition, as all participants completed the study. Tangible resources required for implementation were available. Curriculum content was delivered on slides and displayed using the projector and screen available in the venue. Participants verbally expressed concerns about workshop length and fatigue during the session. To remain within the allotted four-hour timeframe, two of the original ten prompts were skipped, and the facilitator quickly progressed through modules 8 and 9. The completeness of the survey data differed by domain, with each participant completing items in the engagement and impact domains; however, seven and nine participants completed all trust and humor domains, respectively (Table 3).

**Table 3:** Pre- and Post-Intervention Descriptive Summary Statistics.

| Table 3: Pre- and Post- Intervention Descriptive Summary Statistics |  |  |  |  |  |  |  |
| --- | --- | --- | --- | --- | --- | --- | --- |
| Domain | N | Pre-Intervention |  |  | Post-Intervention |  |  |
|  |  | Mean | SD | Range | Mean | SD | Range |
| Trust | 7 | 3.1 | 0.7 | 1.92–4.38 | 3.0 | 0.8 | 1.62–4.15 |
| Engagement | 10 | 4.0 | 0.7 | 3.33–5 | 4.2 | 0.5 | 3.33–5 |
| Impact | 10 | 4.2 | 0.6 | 3.33–5 | 4.3 | 0.7 | 3.33–5 |
| Humor | 9 | 4.7 | 0.4 | 4–5 | 4.5 | 0.6 | 3.5–5 |

### Engagement and Theme Diversity

Results from the rapid thematic analysis were used to derive two engagement measures including response count per prompt, and theme count per prompt. The mean response count was highest during the mid prompt period (9.33; Figure 1A) and lowest during the late prompt period (2.50; Figure 1A). Additionally, there was a 65.9% reduction in the mean total responses between the early and late prompt periods. Mean theme counts followed a similar pattern, with the mid period highest (5.00, Figure 1B) and the late period lowest (2.00, Figure 1B). Linear regression of response count on prompt order yielded a negative association (β_1_ = −0.99), interpreted descriptively given the small number of eight prompts (Figure 1A). The highest individual response count occurred during the fourth prompt, which addressed barriers to successful academic-community partnerships, with 13 responses (Table 2). Free-text responses indicated participant interest in spending more time on the first module, which addressed reasons for scientific and medical mistrust among Black Americans. Furthermore, theme diversity, a metric created to understand variety of themes across prompts, increased across periods (early 0.44, mid 0.53, late 0.75, Figure 1C), indicating that late-period responses contributed proportionally more distinct ideas (Table 2).

### Exploratory Descriptive Analysis of Survey Data

Survey items were categorized into four domains including comfort with humor in scientific communication (Humor), willingness to engage with scientists (Engagement), perception of research as a useful tool for environmental justice (Impact), and trust in scientists (Trust) Following the intervention, descriptive data showed decreases in scores for the Humor and Trust domains and increases in scores for the Engagement and Impact domains (Table 3). Substantial variability across participant responses was observed.

### Qualitative Analysis of Workshop Prompts

The short, free-text responses to each workshop prompt were analyzed using rapid inductive thematic coding, and major themes across prompts related to principles of community-engaged participatory research (Table 2). When asked what comes to mind regarding research partnerships, participants highlighted an approach that includes collaboration, clear division of labor, mutual respect, community initiation, and direct involvement in research tasks. Scientists were seen as bringing valuable resources and specialized knowledge to these partnerships, while community members contribute their own expertise and emphasize local priorities. Barriers to successful partnerships were identified across a range of themes, including perceived bias from scientists, insufficient communication, lack of information, scientific mistrust, fear, unresolved conflicts over funding, and failure to center community interests in research design. Suggestions for a community bill of rights reflected priorities such as addressing tangible needs, fostering shared responsibility, ensuring respect for community contributions, aligning goals from the outset, and maintaining engagement through project completion. Reflecting on previous research experiences, participants described a spectrum of involvement where some studies featured heavy community participation, while others were more limited. Looking to the future, respondents expressed a strong desire for greater involvement, particularly through community-based participatory research. Finally, emerging themes regarding science and epidemiology as tools to support their community include generating understanding and concern, sharing experiences, and enabling community members to learn more about science.

## DISCUSSION

Our feasibility study was delivered as an educational intervention designed for potential academic and community stakeholders initiating a community-engaged research partnership. The intervention met recruitment and attendance targets while identifying areas for refinement, including workshop duration, survey instrument design, and module prioritization, which are being addressed through workshop co-development with a community advisory board (CAB) composed of research participants from this study. Results from this study will inform the pilot study, which will include a larger sample of Altgeld Gardens residents and investigate whether the intervention’s community-level, capacity-building aims produce the intended effects, such as mutual knowledge exchange between community and academic partners, greater scientific trust, more favorable views of research as a vehicle for environmental justice, and increased willingness to collaborate.

Our workshop contributes to a small but growing set of resources available to researchers new to community-engaged research. In the CE Studio model, the flow of information goes from community stakeholders to researchers while a neutral moderator facilitates the interaction [22]. Overall, CE Studio produces input for research projects while minimizing researcher burden by involving a dedicated Community-Engaged Research Core staffed with trained facilitators. Our workshop is designed differently by addressing the partnership-formation phase rather than project-specific consultation. In our model, information flows bidirectionally, and interactions are facilitated directly by the researcher, requiring the development of community-engagement skills. Therefore, direct facilitation is the mechanism through which both academic and community partners build capacity for sustained partnership work.

Feasibility, specifically successful recruitment and workshop delivery, depended on combining community and academic resources. PCR contributed their established recruitment methods that leveraged their social capital in the community and selected the community-based venue for the workshop, both of which supported meeting recruitment targets. On the academic side, the research team secured funding that supported intervention materials, gift card compensation, and lunch. Researchers new to community-engaged work will likely encounter the practice of providing meals during longer sessions for the first time when facilitating this workshop. For participant recruitment, we strongly recommend that genomics researchers work through established community partner channels instead of recruiting individuals independently. The transferability of the partnership model depends on the specific context, including the community, the partnering organization, and the geographic setting. The workshop’s bidirectional structure proved feasible to deliver and produced outputs usable beyond the workshop. Both academic and community partners are positioned as teachers and learners, and the workshop surfaced information about community priorities that were documented and will be carried forward into the partnership. For example, responses to the prompt about a community bill of rights will be used in future memoranda of understanding and governing documents for our community-academic partnership. In addition, information shared throughout the workshop about each stakeholder’s area of expertise can be referenced when co-developing research questions and projects.

Workshop duration emerged as the principal implementation constraint. Participant engagement was strongest during early and middle workshop modules and declined during the final portion of the four-hour session. Verbal feedback during the workshop, observed participant fatigue, and the engagement metrics identified duration as a limiting factor, and time limitations meant that two of the ten planned prompts were not delivered. Although the intervention was originally designed to span four one-hour sessions across separate days, we delivered it as a single four-hour session following the recommendation of a PCR representative who anticipated multi-day retention challenges. The CAB of PCR members is currently co-developing the pilot intervention to shorten the workshop and identify content that can be moved to optional handouts or a companion webpage. Our co-development approach aligns with published community-based participatory research models in which interventions are designed and refined with community partners [39].

Humor was an intentional design element of the workshop and was assessed as one of the four survey domains. Humor was integrated as a pedagogical tool to build community stakeholders’ genomic knowledge and support meaningful engagement with genomics research. Scientist-delivered humor has been associated with increased public trust in scientists; however, studies in this area did not focus on communities historically harmed by scientists or trust-building in a community-engaged research context [31, 32]. In this feasibility study, participants reported lower comfort with humor in scientific communication after the intervention than before. The small sample size, and missing responses make this finding difficult to interpret. The next iteration will include humor co-developed with the CAB and stand-up comedians to better align with community preferences, consistent with findings that inside jokes increase audience laughter during scientific presentations [40].

Survey administration and content both require refinement before the pilot study. Paper-based administration resulted in missing item responses, which reduced the number of participants contributing to domain-level analyses. We plan to use electronic surveys requiring a response before advancing for the pilot study. Participants also reported reduced trust following the intervention. The small sample and the adapted nature of the trust measure limit what can be concluded from this change. The current trust domain measures trust in scientists and science generally. However, partnership formation depends on trust between the community and the specific genomics researcher engaging them. Consequently, we will add an additional domain to the survey assessing trust in the community-engaged researcher, distinct from trust in science as an institution.

Our feasibility study has several limitations beyond those addressed above. Half of participants were PCR members familiar with the workshop content, which may have contributed to engagement during prompts but limits the representativeness of the sample for inferring how the intervention would land with community members new to community-academic research partnerships. While the pilot intervention is being co-developed with the CAB, the present feasibility intervention was developed with limited direct community input beyond informal conversations with PCR. In addition, qualitative coding was conducted by a single researcher without a second coder. These design and analysis limitations are being addressed for the pilot study.

## CONCLUSION

Our feasibility study demonstrated that a humor-integrated, community-engaged educational intervention designed for both academic and community partners can be successfully delivered in partnership with a trusted community organization. The findings identify specific implementation refinements, particularly in workshop duration, survey instrument design, and module prioritization, that are being addressed through ongoing co-development with a CAB. The workshop curriculum, structured prompts, and forthcoming refined survey instrument together constitute a toolkit for genomics researchers without prior community-engaged research training as they initiate sustainable partnerships with Black American communities. The broader goal of this work is 1) to provide academic and community partners with an efficient, structured, and replicable approach for mutually orienting to one another and assessing the suitability of prospective partnerships before committing to long-term collaboration, and 2) to lay the foundation for future community-engaged environmental health research in communities disproportionately burdened by exposure-related health outcomes.

## Data Availability

All data produced in the present study are available upon reasonable request to the authors.

## Author Contributions

LCH conceptualized and designed the study, acquired funding, conducted the analyses, created the figures, and wrote the original draft. KS, MR, KH, and BW contributed to the design of the curriculum and intervention. LCH, KS, and MR developed the survey instrument. LCH, KS, MR, BW, and MT interpreted the data. MT provided supervision. All authors reviewed and edited the manuscript and approved the final version.

## Acknowledgements

The authors want to thank People for Community Recovery and each research participant for contributing to the study.

## REFERENCES

1. Kim, I.E., Jr. and I.N. Sarkar, Racial Representation Disparity of Population-Level Genomic Sequencing Efforts. Stud Health Technol Inform, 2019. 264: p. 974–978.

2. Spratt, D.E., et al., Racial/Ethnic Disparities in Genomic Sequencing. JAMA Oncol, 2016. 2(8): p. 1070–4.

3. Corpas, M., et al., Bridging genomics’ greatest challenge: The diversity gap. Cell Genom, 2025. 5(1): p. 100724.

4. Popejoy, A.B. and S.M. Fullerton, Genomics is failing on diversity. Nature, 2016. 538(7624): p. 161–164.

5. Bentley, A.R., S. Callier, and C.N. Rotimi, Diversity and inclusion in genomic research: why the uneven progress? J Community Genet, 2017. 8(4): p. 255–266.

6. Manrai, A.K., et al., Genetic Misdiagnoses and the Potential for Health Disparities. N Engl J Med, 2016. 375(7): p. 655–65.

7. Need, A.C. and D.B. Goldstein, Next generation disparities in human genomics: concerns and remedies. Trends Genet, 2009. 25(11): p. 489–94.

8. Bustamante, C.D., E.G. Burchard, and F.M. De la Vega, Genomics for the world. Nature, 2011. 475(7355): p. 163–5.

9. Petrovski, S. and D.B. Goldstein, Unequal representation of genetic variation across ancestry groups creates healthcare inequality in the application of precision medicine. Genome Biol, 2016. 17(1): p. 157.

10. Thakur, N., et al., Enhancing Recruitment and Retention of Minority Populations for Clinical Research in Pulmonary, Critical Care, and Sleep Medicine: An Official American Thoracic Society Research Statement. Am J Respir Crit Care Med, 2021. 204(3): p. e26–e50.

11. Washington, H.A., Medical Apartheid: The dark history of medical experimentation on Black Americans from colonial times to the present. 2006, New York: Random House.

12. Scharff, D.P., et al., More than Tuskegee: understanding mistrust about research participation. J Health Care Poor Underserved, 2010. 21(3): p. 879–97.

13. Dye, T., et al., Sociocultural variation in attitudes toward use of genetic information and participation in genetic research by race in the United States: implications for precision medicine. J Am Med Inform Assoc, 2016. 23(4): p. 782–6.

14. Corbie-Smith, G., et al., Attitudes and beliefs of African Americans toward participation in medical research. J Gen Intern Med, 1999. 14(9): p. 537–46.

15. Help this Garden Grow. Respair Production and Media.

16. Smith, L.T., Decolonizing Methodologies: Research and Indigenous Peoples. Third ed. 2021: Bloomsbury Publishing.

17. Wilkins, C.H., et al., Community-Engaged Research - Essential to Addressing Health Inequities. N Engl J Med, 2023. 389(21): p. 1928–1931.

18. Jagosh, J., et al., A realist evaluation of community-based participatory research: partnership synergy, trust building and related ripple effects. BMC Public Health, 2015. 15: p. 725.

19. Israel, B.A., Methods for community-based participatory research for health. 2nd ed. 2013, San Francisco: Jossey-Bass.

20. Khodyakov, D., et al., On using ethical principles of community-engaged research in translational science. Transl Res, 2016. 171: p. 52–62 e1.

21. Lemke, A.A., et al., Addressing underrepresentation in genomics research through community engagement. Am J Hum Genet, 2022. 109(9): p. 1563–1571.

22. Joosten, Y.A., et al., Community Engagement Studios: A Structured Approach to Obtaining Meaningful Input From Stakeholders to Inform Research. Acad Med, 2015. 90(12): p. 1646–50.

23. Genetics, A.S.o.H. ASHG Issues New Guidance Addressing Underrepresentation in Genomics Research Through Community Engagement. 2022 [cited 2026; Available from: https://www.ashg.org/policy/new-guidance-addressing-underrepresentation-in-genomics-research/.

24. White, B.M. and E.S. Hall, Perceptions of environmental health risks among residents in the “Toxic Doughnut”: opportunities for risk screening and community mobilization. BMC Public Health, 2015. 15: p. 1230.

25. Cancer diagnosis rate 2017-2021, based on data from the Illinois State Cancer Registry, available on the Chicago Health Atlas. 2025 [cited 2025; Available from: https://chicagohealthatlas.org

26. Distant/systemic cancer diagnosis rate, based on data from the Illinois State Cancer Registry, available on the Chicago Health Atlas. 2025 [cited 2025; Available from: https://chicagohealthatlas.org

27. Particulate matter (PM 2.5) concentration in 2020, based on data from the U.S. Environmental Protection Agency, available on the Chicago Health Atlas. 2025 [cited 2025; Available from: https://chicagohealthatlas.org

28. Thomas PA, K.D., Hughes MT, Chen BY., Curriculum Development for Medical Education: a Six-Step Approach. Third ed. 2016, Baltimore, MD: Johns Hopkins University Press.

29. Espín-Pérez, A., et al., Short-term transcriptome and microRNAs responses to exposure to different air pollutants in two population studies. Environ Pollut, 2018. 242(Pt A): p. 182–190.

30. Michael Cacciatore, S.Y., Amy Becker, Ashley Anderson, and Kasha Patel, Cultivating Interest in Science Through Humor: Mirth as a Leveler of Gaps in Science Engagement. Environmental Communication, 2025. 19(8): p. 1526–1535

31. Yeo, S.K., et al., Scientists as comedians: The effects of humor on perceptions of scientists and scientific messages. Public Underst Sci, 2020. 29(4): p. 408–418.

32. Alexandra L. Frank, M.A.C., Sara K. Yeo, and Leona Yi-Fan Su, *Wit meets wisdom:* the relationship between satire and anthropomorphic humor on scientists’ likability and legitimacy. JCOM, 2025. 24(1).

33. Chabeli, M., Humor: a pedagogical tool to promote learning. Curationis, 2008. 31(3): p. 51–9.

34. Zhou, W. and J.C. Lee, Teaching and learning with instructional humor: a review of five-decades research and further direction. Front Psychol, 2025. 16: p. 1445362.

35. Yeo, S.K., et al., Examining the Use of Aggressive Satirical Humor on Perceptions of Trustworthiness in Communication About Renewable Energy. Science Communication, 2025: p. 10755470251345746.

36. Berge, M. and P. Anderhag, The Role of Joking for Learning Science: An Exploration of Spontaneous Humour in Two Physics Education Settings. Science & Education, 2025. 34(4): p. 2331–2352.

37. Pascoe, G. and R. Kaplan, Humorous lectures and humorous examples: Some effects upon comprehension and retention. Journal of Educational Psychology, 1977. 69(1): p. 61–65.

38. LaVeist, T.A., L.A. Isaac, and K.P. Williams, Mistrust of health care organizations is associated with underutilization of health services. Health Serv Res, 2009. 44(6): p. 2093–105.

39. Salunke, J., et al., Community Collaboration in Public Health Genetic Literacy: Methods for Co-Designing Educational Resources for Equitable Genomics Research and Practice. Public Health Genomics, 2025. 28(1): p. 66–84.

40. Mammola, S., et al., Statistically significant chuckles: who is using humour at scientific conferences? Proceedings of the Royal Society B: Biological Sciences, 2026. 293(2067).

